# A Highly Generalizable Natural Language Processing Algorithm for the Diagnosis of Pulmonary Embolism from Radiology Reports

**DOI:** 10.1101/2020.10.13.20211961

**Authors:** Jacob Johnson, Grace Qiu, Christine Lamoureux, Jennifer Ngo, Lawrence Ngo

**Affiliations:** CoRead AI, Durham, NC 27712; University of Wisconsin School of Medicine and Public Health, Madison, WI 53726; University of Wisconsin-Madison, Madison, WI 53714; Virtual Radiologic, Eden Prairie, MN 55344; Department of Radiology, Duke University Medical Center, Durham, NC 27710

## Abstract

Though sophisticated algorithms have been developed for the classification of free-text radiology reports for pulmonary embolism (PE), their overall generalizability remains unvalidated given limitations in sample size and data homogeneity. We developed and validated a highly generalizable deep-learning based NLP algorithm for this purpose with data sourced from over 2,000 hospital sites and 500 radiologists. The algorithm achieved an AUCROC of 0.995 on chest angiography studies and 0.994 on non-angiography studies for the presence or absence of PE. The high accuracy achieved on this large and heterogeneous dataset allows for the possibility of application in large multi-center radiology practices as well as for deployment at novel sites without significant degradation in performance.

## Introduction

Pulmonary embolism (PE) is a life-threatening condition that occurs when blood clot from a deep vein thrombosis (DVT) travels into the pulmonary vasculature (1). Although computed tomography pulmonary angiography (CTPA) has become the *de facto* gold standard for workup of PE, diagnosis is challenging because technical factors (e.g., motion artifact and contrast bolus timing) can decrease the confidence of diagnoses. Accordingly, the language in radiologists’ CTPA reports can be quite variable - up to 2296 distinct words have been found to be associated with a diagnosis of PE alone, which significantly increases the complexity associated with developing automated algorithms for the classification of these reports (2).

Earlier approaches to algorithm development used lexical rules to classify medical imaging reports with moderate levels of performance; for example, 0.847 accuracy in one case (3) and an F1 score of 0.867 in another (4,5). Such approaches had difficulty with dealing with certain classes of diagnoses, such as non-PE related thrombosis. Furthermore, they can be quite labor-intensive to develop and may not generalize across different languages and pathologies. Recent approaches have leveraged more sophisticated strategies (including convolutional neural networks) and achieved higher levels of performance with AUCROC of at least 0.97 and above (5–7).

However, the generalizability of these algorithms remains unvalidated. Most of the prior work involves datasets derived from one or a few number of sites (3,5,6). Thus, regional differences in dictation style may exploit undiscovered weaknesses in these algorithms. Prior work has also focused on the development and validation of these algorithms on CTPA reports. A significant number of PEs are diagnosed on non-CTPA protocol examinations (8), so a highly generalizable algorithm should also be validated on these types of studies as well.

In the current study, we develop a highly generalizable NLP algorithm with training data derived from a large number of hospital centers and radiologists as well as CT protocols. The high level of generalizability increases the confidence by which such an algorithm could be implemented at any given site or clinical scenario without dedicated situation-specific training.

## Methods

This project was performed with an IRB approval for waiver of consent by an external IRB, Western IRB. LN and JJ are co-founders in CoRead AI, which provided the computational resources for the project. Deidentified radiology reports were obtained from Virtual Radiologic (vRad), which was sourced from over 2,000 hospital sites and 500 radiologists. An initial set of radiology reports were reviewed by LN, a radiologist with four years of radiology experience and 12 years of general medical experience, with the help of a NLP tool developed in-house based on lexical rules to categorize reports as either “PE present” or “PE absent.” This tool searched radiology reports using a set of regular expressions designed to capture negative reports. In total, this represented 396 unique phrases and variants, which radiologists used to indicate a diagnosis of “no pulmonary embolism.” Such phrases included, “No evidence of pulmonary embolism” or “No gross evidence of central pulmonary artery embolism.” Exams that included any of these phrases were automatically classified as being negative. Exams that did not include any of these phrases were manually reviewed by LN and classified accordingly. More details on the criteria used for this binary classification are presented in the Supplemental Methods.

Once initial classification was performed, labels were compared to those run from a separate image algorithm (9) to look for discrepancies between the two algorithms. Any discrepancy was manually reviewed and reclassified with the appropriate label.

At the end of an iterative process, a total of 17,474 reports were collected from exams performed from March to April 2017. Additional discrepancy data was collected from exams from January to December 2018. The training set consisted of 2,166 positive exams and 12,069 negative exams. The initial test set consisted of 249 positive exams and 2990 negative exams. No preprocessing was performed on any of the reports. Using a deep bidirectional transformer with pre-trained weights (BERT) (10), training was performed, which resulted in an accuracy of 0.99 on the test set. However, since the entirety of the training and test sets had not been manually reviewed, but instead derived from the hybrid NLP review method described above, more comprehensive testing was performed on a dataset that was better characterized. This set included 12,202 reports from August and September of 2019 (please refer to Supplemental Methods for more details).

## Results

Our BERT-based algorithm had an AUROC of 0.995 on angiography studies and 0.994 on non-dedicated CT studies with contrast. It had a sensitivity of 0.965 on angiography studies and 0.954 on non-angiography studies. It had a specificity of 0.990 on angiography studies and 0.991 on non-angiography studies.

## Discussion

We developed a natural language processing algorithm for the classification of PE on radiology reports with state-of-the-art performance compared to the literature. Several components of our approach are novel and may contribute to the high performance of this algorithm. First, a non-deep-learning algorithm was used to assist in the labeling of reports for the training set, given the large number. To supplement this methodology, we further employed the use of an image processing AI algorithm (9). Through the use of this image-based algorithm, we were able to identify studies for which there was a discrepancy between the image algorithm and NLP. In cases where this was due to NLP errors, we were able to add such cases to the training set and repeat training iteratively.

Further, several aspects of the current study demonstrate the generalizability of such an algorithm. First, we did no preprocessing of the reports before training and inference. Previous approaches typically include only impressions (7), and such an approach may be brittle to differences in report formatting and language, depending on the site. For example, some practices may list their impressions first in a report, while others list them at the bottom. Further, some radiologists may label their impression with an alternative name, such as “conclusion” or “opinion.” By including the entire report without any preprocessing, we are able to ensure robustness across such variations. Additionally, training on the entire report allows for the opportunity of looking for within-report discrepancies. As a topic for future study, one could parse a report into the “findings” and “impressions” sections and run inference separately on each of these partitions. Any discrepancies between the two could conceivably represent a report error that may be worth review.

The scope of our dataset also contributes to generalizability. Our dataset is derived from over 2,000 different hospital sites with around 500 different radiologists represented. This allows for a great diversity in clinical presentation and dictation style to be trained into our algorithm, which aids in its robustness if such an algorithm were to be applied to a novel site. Such a level of generalizability is critical for any application that may be applied to a massively distributed practice, like the one from which our data was derived (vRad). Additionally, given the large size of our test set, we were able to stratify the performance of the algorithm for both angiography and non-angiography studies. This is important to validate because it has previously been shown that non-PE thrombotic disease, such as portal vein thrombosis, may serve as confounds for a PE NLP algorithm (3). Such pathologies are not typically captured on CTPA examinations, and the algorithm’s high accuracy on non-CTPA exams suggests that such an NLP algorithm would be useful and robust for a variety of clinical uses for non-CTPA exams.

Several limitations in our current study lend possible directions for future study. First, our chest angiography cases were not explicitly divided into CTPA exams and non-CTPA angiography cases. The reason for this is that our current dataset does not include labels or tags for which type of CT protocol was used for each of these studies. An additional complicating factor is that contrast-bolus timing can be highly variable from patient to patient, such that an intended CTPA may end up being timed for the aorta or vice versa. This means that some small proportion of chest angiography cases may have been optimally timed for the detection of aortic pathology instead of pulmonary artery pathology. However, given that our performance remains high on non-angiography cases, we have reason to believe that there may not be a significant difference in performance between CTPA and non-CTPA chest angiography cases. A future direction of study could be the development of an image algorithm to delineate the difference between CTPA and non-CTPA angiography, which inform this type of sub-analysis.

Another potential confound is that vRad’s reporting system is mostly structured. Although there is not 100% adherence, the vast majority of vRad’s reports are structured, which may weaken confidence in out-of-sample generalizability. However, when the reports are split into findings and impressions and trained as separate data samples, we retain a high validation accuracy of 0.991, which is similar to the validation accuracy that we report for the whole report training. Since the impression is not structured, this supports the fact that our algorithm would be robust to non-structured reporting as well. Future studies using our algorithm to validate its accuracy on non-structured reports could provide further support.

Finally, we have seen that the development of sophisticated NLP can often require large datasets (11). As far as we are aware, our current dataset of around 30,000 reports represents the largest dataset used for these purposes of which we are aware. This ensures not only that the performance is high, but that the model is robust to heterogeneity in reports that may come from a variety of sources.

The development of a highly generalizable NLP algorithm for PE opens the opportunity for a variety of large-scale studies in the future. For example, the comparison of the results of an image algorithm and NLP output could be useful in the detection of radiologist errors in interpretation. Further, as mentioned above, higher degrees of sophistication in the training and inference of these NLP models on separate sections of radiologist reports could assist in identifying dictation errors.

## Data Availability

The data from this study is not publically available.

## Table

**Table 1:** Performance of an NLP algorithm for classification for presence or absence of PE on angiography and non-angiography reports.

|  | CT Angiography | Non-angiography CT |
| --- | --- | --- |
| AUROC | 0.995 | 0.994 |
| Sensitivity | 0.965 | 0.954 |
| Specificity | 0.990 | 0.991 |

## Figure

**Figure 1:**
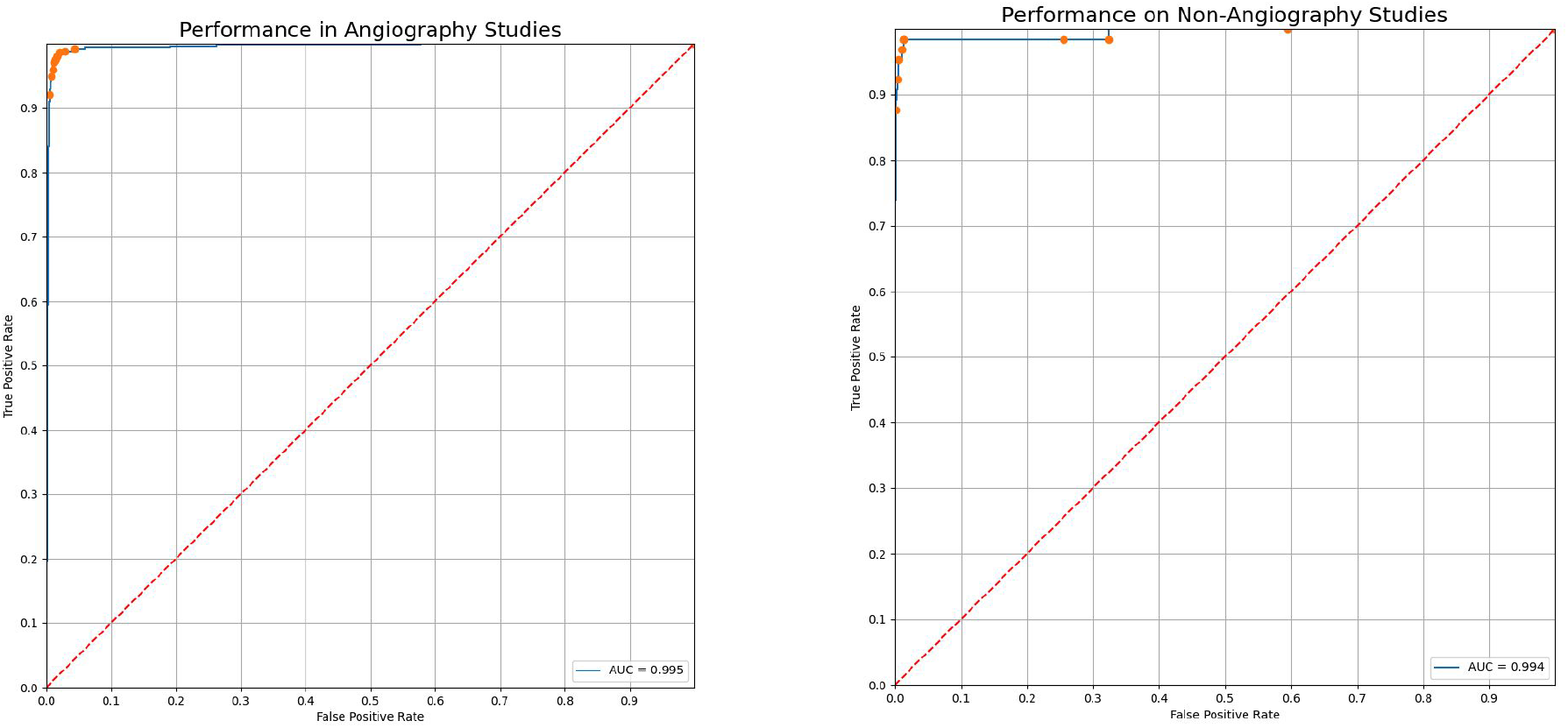
Performance of an NLP algorithm for classification for presence or absence of PE on angiography (AUC 0.995) and non-angiography reports (AUC 0.994).

## Supplemental Methods

### Test set curation

The following is excerpted from previous work on an imaging algorithm (9) and written verbatim for reference:

An additional 12,202 unique CT examinations, which cover the chest anatomy, between July and September 2019 were collected for the test set. This was randomly sampled from a larger set of CT examinations that were collected within the vRad practice. The test set was 51.3% female with a mean age of 54.2. Of these, 7,462 cases were dedicated angiography cases while 4,740 cases were non-dedicated angiography cases. Data in this study were obtained across the United States from more than 2,000 hospital sites, representing nearly all brands and models of CT scanners. The majority of scans were obtained in the emergency department setting with the remainder obtained from inpatients and outpatients.

The report for each examination in the test set was manually reviewed by one of two radiologists, LN or JN, for whether the examination was positive or negative. They have 4 and 7 years of experience in radiology, respectively. At the time of report labeling, they were both blinded to the results of any of the relevant algorithms for any of these exams. Given differences in confidence conveyed by the interpreting radiologists in these reports, rules were established to determine how to perform such a binary classification. The following were labeled as positive exams: (1) clear diagnoses of pulmonary embolism (e.g., “Positive for pulmonary embolism in the right lower lobe.”), (2) chronic pulmonary embolism, (3) a focal finding is questioned without an alternative diagnosis being favored (e.g., “Questionable superior segment right lower lobe pulmonary embolism.”).

The following were labeled as negative exams: (1) clear diagnosis of no pulmonary embolism (e.g., “No pulmonary embolism.”), (2) general limitation of the study (e.g. “No evidence of pulmonary embolus to the segmental level” or “Nondiagnostic study for pulmonary embolism.”), (3) an alternative diagnosis for a focal finding seems to be higher in probability (e.g. “The left lower lobe pulmonary artery contrast bolus fades out in the left lung base. May be related to impeded flow in the atelectatic lung; however, pulmonary embolism cannot be excluded.”).

## Acknowledgements

We would like to thank Brian Baker and his team at vRad for their support of this project.

## Notes

### Competing Interest Statement

Lawrence Ngo and Jacob Johnson are Co-Founders of CoRead AI.
Lawrence Ngo and Chrisitine Lamoureux work as radiologists for Virtual Radiologic.

### Funding Statement

No external funding was provided for this study.

### Author Declarations

This study was approved with a waiver of consent by Western IRB.

